# Domain Knowledge Augmented Contrastive Learning on Dynamic Hypergraphs for Improved Health Risk Prediction

**DOI:** 10.1101/2025.01.18.25320096

**Authors:** Akash Choudhuri, Hieu Vu, Kishlay Jha, Bijaya Adhikari

## Abstract

Accurate health risk prediction is crucial for making informed clinical decisions and assessing the appropriate allocation of medical resources. While recent deep learning based approaches have shown great promise in risk prediction, they primarily focus on modeling the sequential information in Electronic Health Records (EHRs) and fail to leverage the rich mobility interactions among health entities. As a result, the existing approaches yield unsatisfactory performance in downstream risk prediction tasks, especially tasks such as *Clostridioides difficile* Infection (CDI) incidence prediction that are primarily spread through mobility interactions. To address this issue, we propose a new approach that leverages Hypergraphs to explicitly model mobility interactions to improve predictive performance in health risk prediction tasks. Unlike regular graphs that are limited to modeling pairwise relationships, hypergraphs can effectively characterize the complex high-order semantic relationships between health entities. Moreover, we introduce a new contrastive learning strategy that exploits the domain knowledge to generate semantically meaningful positive (homologous) and negative (heterologous) pairs needed for contrastive learning. This unique contrastive pair augmentation strategy boosts the power of contrastive learning by generating feature representations that are both robust and well-aligned with the domain knowledge. Experiments on two real-world datasets demonstrate the advantage of our approach in both short-term and long-term risk prediction tasks, such as *Clostridioides difficile* infection incidence prediction and MICU transfer prediction. Our framework obtains gains in performance up to 29.49 % for PHOP, 30.64 % for MIMIC-IV for MICU transfer prediction, 13.17 % for PHOP, and 4.45 % for MIMIC-IV for CDI Incidence Prediction.

## 1 Introduction

Health risk prediction tasks such as in-hospital mortality prediction [1], likelihood of adverse complications [2], and predicting the onset of healthcare-associated infections (e.g., *Clostridioides difficile* Infection [3]) are crucial for making informed clinical decisions. Recent advances in deep learning based risk prediction models [4–6] have shown great promise in leveraging the longitudinal information present in Electronic Health Records (EHR) and forecasting the health status of patients. For example, [7] proposed Recurrent Neural Network (RNN) based approach to model the temporal visit-level information present in EHR for risk prediction. Similarly, [5] and [8] proposed attention networks to accurately capture the latent information embedded in the hierarchy of visits for health risk prediction.

Despite significant advances, the existing approaches mainly focus on modeling the sequential information present in EHRs and fail to leverage the *mobility interactions* (e.g., patient-doctor, patient-room, and patient-medication) among health entities. This leads to limited representational power for existing approaches, especially for tasks such as *Clostridioides difficile* Infection (CDI) incidence prediction that are primarily spread through mobility interactions. To address this, more recently graph neural networks (GNNs [9]) have been proposed to model the interactions between health entities over graph structures constructed from EHR data. However, GNNs only consider the pairwise interactions which is insufficient to effectively characterize the complex high-order mobility interactions between health entities. For instance, consider a patient *p* that interacts with a healthcare entity *e* at a timestamp *t*. In this scenario, the patient *p*’s aggregated feature representations should be influenced by the features of all the other patients with whom the healthcare entity *e* interacts at time *t*. The rationale being that if a cohort of patients share a room and one of them becomes infected, it is likely that the remaining patients will be infected in the future time. Effectively capturing such high-order mobility interactions that go beyond pairwise relationships is critical for accurately predicting the risk of patients to acquire healthcare-associated infections such as CDI.

To this end, we propose a new approach that explicitly models the higher-order relations between health entities using better suited data structures such as hypergraphs. Hypergraphs offer a more expressive structure than traditional graphs that allow edges to connect more than two nodes (or entities), thus enabling them to encode the semantic relationships between health entities at a granular level. Central to our approach is the proposed hypergraph information aggregation module that aggregates comprehensive information from high-order mobility interactions and learns robust feature representations useful for health risk prediction. Moreover, to further enhance the expressive power of hypergraph modeling, we propose a novel contrastive learning based strategy that enriches the feature representations of health entities by exploiting the prior domain knowledge derived from EHRs. Specifically, the domain knowledge utilized in this paper are: (a) the hierarchical structure of medication codes [10], and (b) the speciality information of doctor, i.e., if two doctors have the same speciality (e.g., infectious disease, cardiology) then they have a semantic relationship [11]. By contrasting the domain knowledge augmented homologous and heterologous pairs, the model learns to generate health entity representations that are expressive, robust, and well-aligned with the prior domain knowledge. Finally, we propose a temporal information aggregation module that aggregates spatial information from previous timestamps, thus enabling the model to adapt to the patient conditions changing over time. Experiments conducted on multiple datasets show that the proposed approach significantly improves the performance in health risk prediction tasks as compared to the baseline methods. In this research, our contributions can be summarized as follows:

- We propose a novel approach that leverages hypergraphs to capture the high order mobility interactions present in EHRs and improves the accuracy for risk prediction tasks. This is different from existing approaches that are primarily focused on modeling the sequential information in EHRs.
- We propose a new contrastive pair augmentation strategy that exploits the domain knowledge to generate semantically meaningful augmentations needed for contrastive learning. This technique enhances the power of contrastive learning for health risk prediction by learning feature representations that are not only robust but also well-aligned with the domain knowledge.
- The proposed approach is evaluated on real-world datasets for both short-term and long-term prediction tasks, including CDI Incidence Prediction and MICU Transfer Prediction. Experimental results demonstrate the effectiveness of the proposed framework in accurately estimating patient risks over various time intervals.

## 2 Methodology

We consider healthcare operations data with events derived from electronic health records (EHRs) [12–15] and admission-discharge-transfer (ADT) records [3, 14–16] from an inpatient healthcare facility. The events in the data log different types of interactions between patients 𝒫 and other entities encountered in healthcare settings, including doctors 𝒟 (visiting patient’s room and/or performing a procedure on the patient), medications ℳ (patient being prescribed a medication) and hospital rooms ℛ (patient being admitted to a room). Each event (*p, e, t, θ*) is an ordered set of four elements which represents interaction between a patient *p* ∈𝒫 and an entity *e* ∈𝒟 ⋃ ℳ ⋃ ℛ at time *t* and of type *θ*. Note that *θ* is an element of the set {‘Doctor’, ‘Medication’, ‘Room’] indicating the type of entity *e*. Without the loss of generality, we assume the events in the data are timestamped between time 0 and some *T >* 0. In addition to the events, healthcare operations data also include demographic and medical information on each patient *p*. These additional personal-level data are temporally dynamic in nature as they include records of health-risk factors such as length of hospital stay, cumulative antibiotic dose count, and gastric acid suppressors use which evolve over the course of patients’ hospital stay. At each timestamp *t*, we represent these data as a feature matrix **X**^*t*^. The *p*^th^ row (i.e.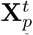) of the matrix represents the feature vector associated with patient *p*.

For a subset of the patients 𝒫_*l*_, we are also given time-stamped log ℒ of lab results (example, CDI test) or decisions made by healthcare providers (example, transfer to MICU) indicating an occurrence of a potentially adverse event. Each entry in ℒ is a tuple (*p, t, l*) indicating that the patient *p* had a log entry at time *t* of type *l*, where *l* is a binary variable that represents whether the outcome was positive or negative. Our main goal in this paper is to utilize all aforementioned data to estimate the risk of adverse events in future timestamps for the remaining patients 𝒫 ∖ 𝒫_*l*_. Figure 1 shows an overview of the proposed approach.

**Figure 1.**
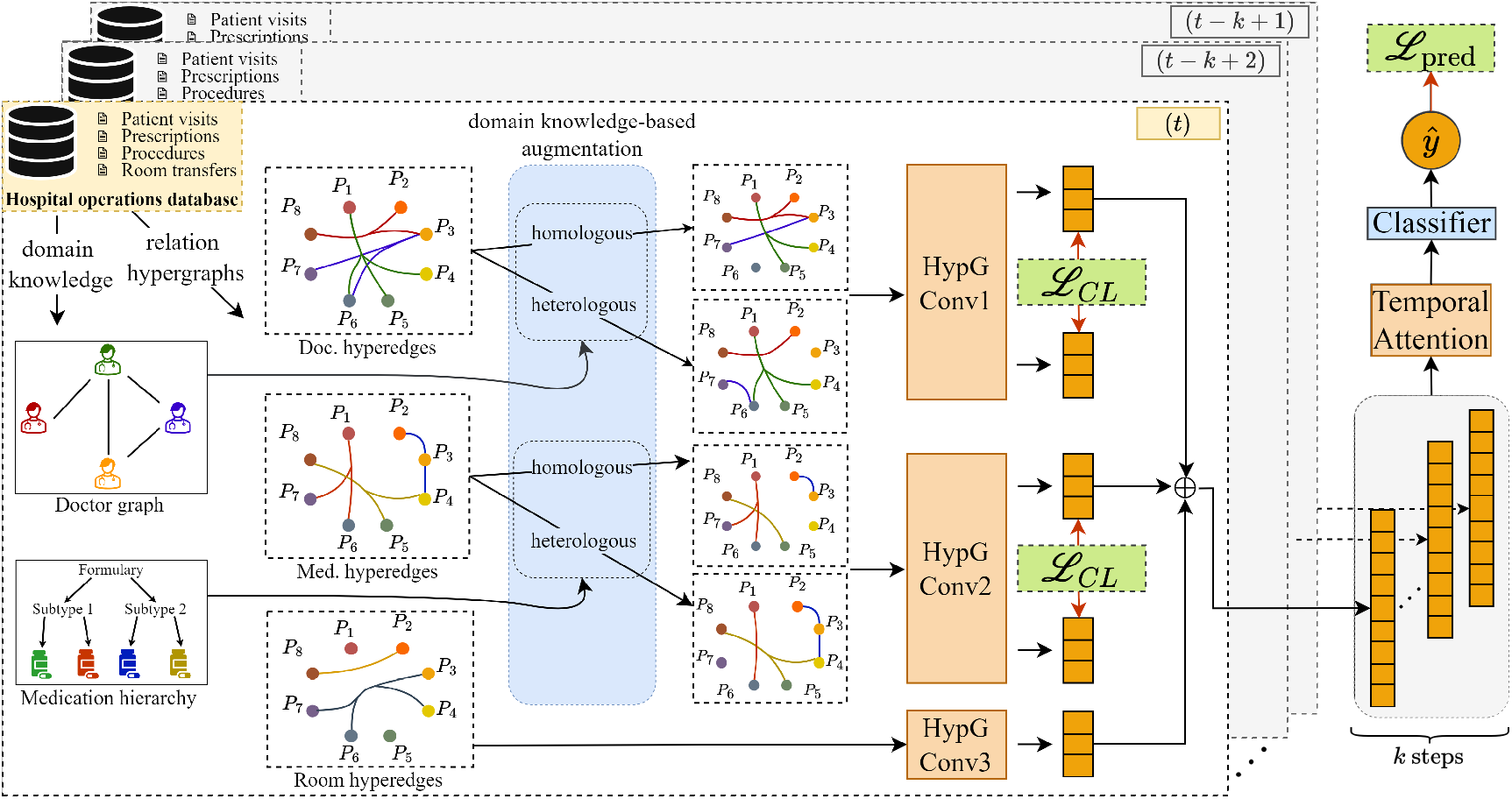
Proposed framework (best viewed in color). We construct domain knowledge capturing relationships between doctors and medications and hypergraphs capturing relationships between patients at each timestamp from dynamic stream of hospital operations data. We construct robust hypergraph augmentations by infusing information from the domain graphs. We then perform patient-hyperedge and hyperedge-patient message passing on both the augmentations and concatenate resultant embeddings from different entity types. We then minimize the contrastive loss between these two augmentations and temporally aggregate the embeddings to make predictions.

### 2.1 Domain Knowledge

Most existing approaches [5, 17] for patient risk estimation do not explicitly utilize the domain knowledge inherently present in the EHR data that goes beyond the patient level. This is limiting because such information could play a critical role in accurate patient risk estimation. For example, two patients examined by two separate cardiologists could have similar risks for cardiovascular diseases. However, this similarity is not captured by the patients’ interactions with the doctors alone. In contrast, it becomes apparent once the relationship between the doctors is taken into account. To address this gap, we propose to extract domain knowledge inherent in EHR data, which describes relationships between entities of a specific type as a static graph

#### *G*_entitytype_ as follows

1. **Doctor Graph:** We construct *G*_doctor_ (𝒟, *E*_doctor_) based on the proximity of the specialty of the doctors with each other. Two doctors *d*_1_ ∈ 𝒟and *d*_2_ ∈ 𝒟 have an edge in *G*_doctor_ if they belong to the same (or similar) specialty.
2. **Medication Graph:** Similarly, we construct *G*_medication_(ℳ ⋃ *M, E*_medication_) as a tree to capture the medication hierarchy. Note that the medications ℳ form the leaf nodes in *G*_medication_ while the intermediate nodes *M* represent medication subtypes.

While our proposed approach leverages the aforementioned graphs, it is generalizable to any domain knowledge expressed as a static graph.

### 2.2 Dynamic Heterogeneous Hypergraph Construction

A hypergraph is usually denoted as *H* = (*V, E*) where *V* denotes the set of the nodes (similar to traditional graphs) and *E* denotes the set of hyperedges that represent relationships between two or more nodes. We use this notion to define our dynamic heterogeneous hypergraph ℌ= {*H*^1^, *H*^2^, …, *H*^*T*^]for *T* snapshots.

Each heterogeneous hypergraph *H*^*t*^ is constructed from interactions occurring at time *t* and is defined as 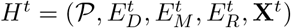where 𝒫 and **X**^*t*^ are the set of patients and feature matrices at time *t* (as defined earlier). 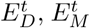 and 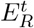 are hyperedges (among patients) of type ‘Doctor’, ‘Medication’, and ‘Room’ respectively. The hyperedges 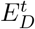 are constructed from the set of interactions *{*(*p, e, t*^*′*^, *θ*)|*t*^*′*^ = *t & θ* = ‘Doctor’]. Each hyperedge 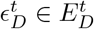 at time *t* connects a subset of patients 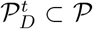 who all have interacted with the same doctor *d* at time *t*, i.e., 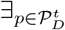(*p, d, t*, ‘Doctor’). The remaining sets of hyperedges 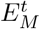 and 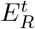 are constructed leveraging the interactions of type ‘Medication’ and ‘Room’ respectively in a similar manner. Also note that each node *p* in subset 𝒫_*l*_ *⊂* 𝒫 (nodes with labels) which has an entry (*p, t, l*) ∈ℒ are labelled as 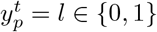 at *t*. Let 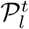 be the set of nodes labeled at time *t*.

Having constructed a partially labeled dynamic heterogeneous hypergraph from hospital operations data, we can now pose patient risk estimation as a semisupervised binary classification problem. As our primary goal is to estimate patient risks in the future, we partition the training and testing data temporally. Let us assume, we train on data until time 0 *< τ < T*, then state our learning problem:

**Given**

A dynamic heterogeneous hyper-graph ℌ= *{H*^1^, *H*^2^, …, *H*^*τ*^ ] and a set of labelled nodes 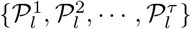

**Infer**

A mapping function *f* (*p*) which maps patients in 𝒫 to a number in the range (0, 1) representing the patient risk of adverse event

**Such that**

a loss function 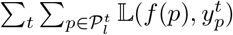 is minimized.

#### 2.2.1 Domain Knowledge Augmented Contrastive Learning

Contrastive learning over graphs has achieved state-of-the-art performance in numerous node classification tasks. However, most prior work on contrastive learning over graphs [18–20] perturb the input graphs by randomly deleting nodes, edges, or features. For example, [20] proposes contrastive learning on hypergraphs deletes the hyperedges and perturbs the node features via a Bernoulli distributed masking matrix. Removing entire hyperedges randomly may introduce unnecessary harmful noise, leading to poor performance (as evidenced by our experiments). To address this issue, we propose to leverage domain information encoded in the graphs *G*_doctor_(which captures doctordoctor similarity) and *G*_medication_ (which represents the medication hierarchy) to augment the hypergraphs by perturbing the structure of the hyperedge instead of completely deleting it. For each graph, we generate both the homologous and heterologous augmentations. To generate homologous augmentation of the heterogenous hypergraph at time *t* based on *G*_doctor_, we iterate through all the edges (*d*_1_, *d*_2_) in *G*_doctor_ keeping track of patients who interact with both *d*_1_ and *d*_2_. We then remove the interactions between such patients and both *d*_1_ and *d*_2_ in our hypergraph at time *t*. The pseudo-code is presented in Algorithm 1. Similarly, to obtain the heterologous augmentation, we first sample a complement of *G*_doctor_ and use it as the Semantic graph input of Algorithm 1. Note that this approach removes the interactions between doctors and patients for patients who interact with doctors who are not connected in *G*_doctor_. We use a similar approach to generate homologous and heterologous augmentations based on *G*_medication_. Here instead of determining the medication-patients relations to be deleted based on the edges of *G*_medication_, we determine it based on the whether two medication belong to the same sub-type.

Having obtained the homologous augmentation 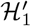 and the heterologous augmentation 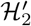, we obtain patient embeddings 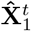 and 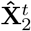 via any standard hy-pergraph convolution step, whose information aggregation steps can be further divided into patient-to-hyperedge and hyperedge-to-patient information aggregation steps.

##### Algorithm 1

Homologous Augmentation

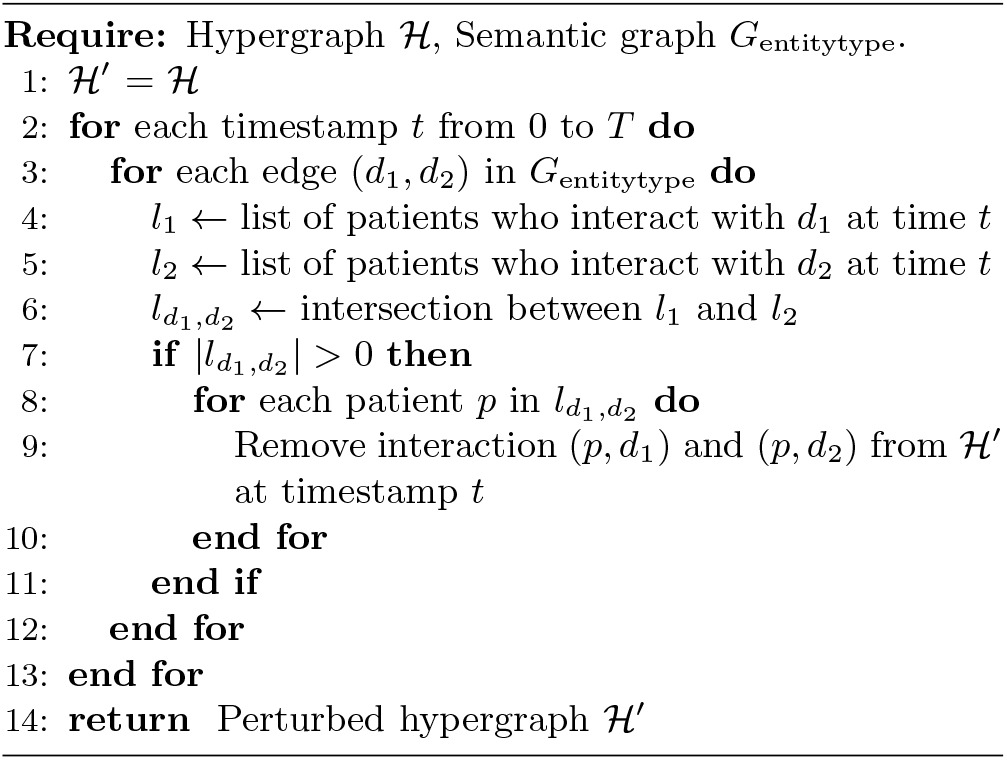

Based on the dynamic heterogeneous hypergraph defined above, we construct hypergraph ℌ convolution for each interaction type e ‘Doctor’, ‘Medication’, and ‘Room’ respectively. For the hyperedges *E*^*t*^ constructed at timestamp *t*, we construct the incidence matrix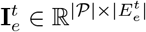, node diagonal matrix 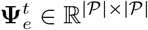 column diagonal matrix 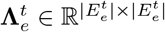. For any entity type *e D, M, R* denoting ‘Doctor’, ‘Medication’, and ‘Room’, the patient-hyperedge aggregation scheme is:

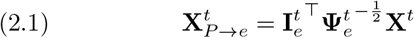

Similarly, the hyperedge-to-patient message aggregation to update the patient representations with the learned hyperedge embeddings is:

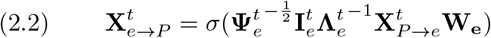

Here,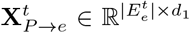 denotes the aggregated hyperedge embedding, 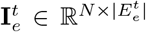 and 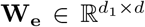 and 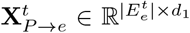 . *σ* is a nonlinear activation function like ReLU, and 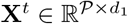. Also 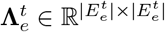.

Then, the deviation between the two augmentations is minimized by the contrastive loss ℒ_*CL*_, which maximizes the similarity between the learned representation of a patient across two augmentations while making these dissimilar from a randomly sampled patient’s representation. The temperature-scaled contrastive loss (NT-Xent) ℒ_*CL*_ is given by:

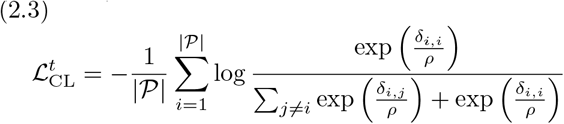

Here *δ*_*i*,*j*_ denotes the cosine similarity between contrastive embedding pairs 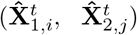 and *ρ* is a temperature-scaling parameter.

Based on the three aggregated patient features, the overall patient feature representation is given by the concatenation operation as shown below:

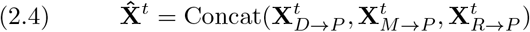

#### 2.2.2 Temporal Aggregation

After obtaining the robust latent patient embeddings from the contrastive learning module, we combine the embeddings learned from the current and prior timestamps to obtain a final representation for each patient. For a patient *p* at timestamp *t*, we select the embeddings of the current timestamp as well as of previous *k* timestamps, which we denote using 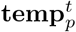. Note that 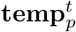 is of size *k ×* 3*d*, where *d* is the output size of our hypergraph convolution network. We then use the weighted sum of the embeddings in this set to obtain the final representation for the patient.

We do this by passing all the *k* embeddings through a Transformer and then using the self-attention mechanism [21] to learn the weights and the final embedding.

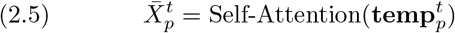

Previous works [1] have identified that information from 2-5 days before an adverse event is useful in predicting the risk of the event occurrence. Empirically, we noticed that *k* = 3 leads to the best performance in our tasks.

#### 2.2.3 Training

After obtaining the updated latent feature representation for patient *p*,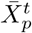 at timestamp t, we obtain the predicted label 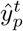 for the patient by passing the feature representation through a Feed-Forward Neural Network (FFN). We minimize the binary cross entropy loss function with the true label 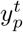 for that patient (if present) which is formulated as:

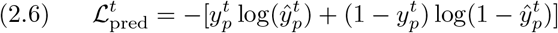

We jointly minimize ℒ_*CL*_ at each timestamp as well as ℒ_pred_ after obtaining the prediction logits via the Temporal Aggregation Module and the Decoder Module. So, our overall objective function to be minimized can be written as:

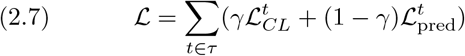

Here *γ*is a hyperparameter. In practice, we aggregate all the patient predictions across all timestamps and use a mask to filter out the patients without labels. Then we compute the binary cross-entropy loss function for all the patients with available labels.

## 3 EXPERIMENTS

### 3.1 Datasets

In our experiments, we used Electronic Healthcare Records (EHR) and Admission Discharge Transfer (ADT) data from 2 real-world healthcare facilities. Statistics of the datasets used for the experiments are given in Table 1.

**Table 1:** Number of interactions in our datasets.

| Interactions | PHOP | MIMIC-IV |
| --- | --- | --- |
| Patient-Doctor | 23,085 | 8,046 |
| Patient-Medication | 349,345 | 34,857 |
| Patient-Room/Unit | 16,771 | 3,334 |

#### PHOP

Proprietary Hospital OPerations data (**PHOP**) is derived from real-time healthcare operations data collected from a large (800-bed) tertiary care teaching hospital in the U.S. mid-western region.

#### MIMIC-IV

The second dataset is derived from the open-source MIMIC-IV data set [29], which contains information on vital signs, prescribed medications, laboratory tests, and procedural events of ICU patients admitted to Beth Israel Deaconess Medical Center (BIDMC).

Further details about the datasets are given in A.1. We conducted all experiments on AMD EPYC 7763 64-Core Processor with 1.08 TB memory and 8 NVIDIA A40 GPUs with CUDA version 12.2.

### 3.2 Baselines

We compare our method against several state-of-the-art baselines. Some of our baselines were designed to work in the static setting, hence we have modified these to make predictions in the dynamic setting for a fair comparison. Approaches like [8, 30] heavily depend on prior hospitalization information (which we do not consider in the current problem setup) and hence cannot be used for fair comparison against our approach. The baselines we contrast our approach are as follows:

#### 1) Patient Feature Baselines

We use domaininspired feature representations encapsulating each patient’s length of stay, history of previous visits, antibiotics prescription, etc. as features for these models. The baseline models are **Logistic Regression** [22] (**LR**) and **Feed-Forward Network** [23] (**FFN**).

#### 2) Domain-Inspired baselines

The **Multitask** Method [26] (**MT**) constructs additional features using the length-of-stay of the patient in addition to the domain-inspired features and trains the model in a multi-task manner. On the other hand, **HypEHR** [28] and **CACHE** [17] use medical codes to construct static hypergraphs to perform predictions. **DECent** [27] is an unsupervised learning method that also incorporates heterogeneous interactions.

#### 3) Sequential Deep Learning Baseline

This approach temporally aggregates information from previous timestamps to make predictions for the current timestamp. we choose **LSTM** [24] to be the baseline here.

#### 4) Deep Learning Baselines Incorporating Interactions

These models leverage the information from patient interactions with hospital entities. We use **Spatiotemporal Graph Convolution Network** (**SGCN**), a modification of the original GCN model [25] with temporal aggregation via self-attention.

#### 3) Contrastive Learning Baseline

**HyGCL** [20] uses contrastive learning by augmenting views on a hypergraph. Unlike our approach, **HyGCL** randomly drops hyper-edges as augmentation.

### 3.3 Tasks

To demonstrate the benefit of using our method for patient risk estimation, we compare our framework and the baselines on two tasks

1. **MICU Transfer Prediction:** The task here is to predict if a patient is at risk of being transferred to a medical intensive care unit (MICU) [14, 15]. Patients are likely to be transferred to the MICU when their conditions deteriorate and require urgent treatment and continuous observation. We pose MICU transfer prediction as a binary classification task in which inpatients transferred to the MICU at particular timestamps are positive instances. The remaining patients are negative instances. All methods need to correctly predict the labels of nodes at time *t*, only using the embeddings learned at or before time *t* − *d*, where *d* is the delay period.
2. **CDI Incidence Prediction:** *Clostridioides difficile* Infection (CDI) is a Hospital Acquired Infection (HAI) that primarily spreads within healthcare facilities [31]. It spreads through direct contact between individuals and also via surfaces (such as the doorknobs in patients’ rooms). Early diagnosis of patients at risk for CDI provides healthcare professionals with a crucial time to fight the disease. Typically a patient is screened for CDI **three days** after exhibiting symptoms. This indicates that the infection might have proliferated before the positive test result [14,15,32]. Hence, motivated by prior works [15, 27, 33], we make 3-day ahead predictions. We pose it as a binary classification problem.

### 3.4 Data Split and Performance Metric: Contrary to the train: test split done in the downstream prediction tasks by [14, 15], we employ a temporal split to define the train: test data and labels to prevent information leakage. We consider the data and labels during 1/1/2010 - 3/31/2010 for **PHOP** and 1/1/2128 - 4/10/2128 (date-shifted true values in the data) for **MIMIC-IV**. Further details are in the appendix

Both the tasks described have highly imbalanced label distribution. MICU Transfer prediction task has about 1:75 label imbalance for **PHOP** and about 1:60 for **MIMIC-IV**. On the other hand, the CDI Incidence prediction task has about 1:88 label imbalance for **PHOP** and about 1:76 for **MIMIC-IV**. The extreme label imbalance makes Area under Receiver Operating Curve (AUROC) a better evaluation metric over accuracy like prior works [26, 27]. For rigorous evaluation, we also report Area under the Precision-Recall Curve (AUPRC) as an additional metric.

### 3.5 Short and Long Term Risk Prediction

#### MICU Transfer

We perform 1 to 3 days ahead MICU Transfer Prediction for both datasets. The results of the experiments are present in Tables 2 & 3. For this task, 1 and 2-day-ahead predictions are shortterm predictions while 3-day-ahead predictions correspond to long-term prediction task. We note that standard baselines that only take individual risk factors (features) into account such as **LR, FFN**, and **MT** typically perform the worst as they do not take temporal, relational, and semantic information into account. Note that **LSTM** which only uses temporal information has a slightly better performance. **SGCN** and **DECent** perform better than **LSTM** as they leverage both interaction and temporal information. Hypergraphs-based methods, **HyGCL, CACHE**, and **HypEHR**, have a strong performance over other baselines indicating the importance of modeling the higher-order relations for the task. We note that our method outperforms **HyGCL** (the only other contrastive-learning baseline) for all the tasks. This means that the infusion of domain knowledge to construct the contrastive pairs improves the overall predictive performance. Our framework obtains gains in performance up to 3.59 % for **PHOP** and 6.02 % for **MIMIC-IV** over the nearest baseline in terms of AUROC and gains in performance up to 29.49 % for **PHOP** and 30.64 % for **MIMIC-IV** over the nearest baseline in terms of AUPRC. We also note that our method significantly outperforms the contrastive-learning baseline **HyGCL** across all tasks, thus highlighting the importance of incorporating domain-inspired augmentations. **CDI Incidence**: We perform 3-day-ahead CDI Incidence Prediction for both datasets. The results of the experiments are presented in Table 4. Similar to the previous experiment, we notice that **LR** and **FFN** perform poorly on both datasets. However, we notice that **MT**, which is designed for this predictive task, outperforms **LR**,**FFN** and **LSTM** in terms of AUROC. As in MICU transfer prediction, the hypergraphs-based approaches perform the best over all baselines while the mobility-interactions-based baselines **DECent** and **SGCN** particularly outperform the static and sequential baselines. We observe that our framework again outperforms all the baselines across all the tasks in terms of AUROC on both datasets highlighting the consistent superiority of our proposed approach over the baselines. We also note our framework has improvements of 3.59 % for **PHOP** and 2.79 % for **MIMIC-IV** over the nearest baseline in terms of AUROC and gains in performance up to 13.17 % for **PHOP** and 4.45 % for **MIMIC-IV** over the nearest baseline in terms of AUPRC. Note that the large difference in AUPRC scores over the two datasets is due to the difference in the number of interactions in the two datasets as shown in Table 1 and as CDI is a disease that spreads through contact, the reduction in the number of pathways of disease transmission makes prediction for **MIMIC-IV**.

**Table 2:** Mean AUROC scores (in percentage) for MICU Transfer Prediction on **PHOP** and **MIMIC-IV** Datasets across 3 runs.

| Model | PHOP |  |  | MIMIC-IV |  |  |
| --- | --- | --- | --- | --- | --- | --- |
|  | 1-day | 2-day | 3-day | 1-day | 2-day | 3-day |
| **LR** [22] | 64.29 ± 0.30 | 71.57 ± 0.21 | 71.73 ± 0.11 | 60.29 ± 0.43 | 61.45 ± 0.17 | 60.18 ± 0.43 |
| **FFN** [23] | 68.26 ± 0.11 | 77.26 ± 0.55 | 72.48 ± 0.14 | 61.66 ± 0.17 | 63.73 ± 0.85 | 61.16 ± 0.29 |
| **LSTM** [24] | 75.18 ± 0.62 | 82.59 ± 0.31 | 82.11 ± 0.73 | 68.65 ± 0.27 | 69.75 ± 0.21 | 68.42 ± 0.34 |
| **SGCN** [25] | 79.18 ± 0.36 | 80.91 ± 0.16 | 85.37 ± 0.52 | 71.26 ± 0.28 | 77.93 ± 0.58 | 73.61 ± 0.49 |
| **MT** [26] | 60.72 ± 0.52 | 70.19 ± 0.16 | 65.98 ± 0.71 | 58.26 ± 0.20 | 54.90 ± 0.11 | 53.83 ± 0.19 |
| **HyGCL** [20] | 87.63 ± 1.14 | 86.61 ± 0.60 | 87.06 ± 0.33 | 80.32 ± 0.59 | 78.40 ± 0.10 | 77.26 ± 0.15 |
| **DECent** [27] | 84.68 ± 0.55 | 75.28 ± 0.19 | 74.93 ± 0.88 | 81.67 ± 1.40 | 77.42 ± 0.83 | 78.35 ± 0.60 |
| **HypEHR** [28] | 87.49 ± 0.21 | 72.36 ± 0.18 | 73.38 ± 0.95 | 65.49 ± 0.34 | 63.67 ± 0.41 | 61.55 ± 0.20 |
| **CACHE** [17] | 89.85 ± 0.35 | 80.54 ± 0.55 | 79.62 ± 0.44 | 79.83 ± 0.37 | 71.21 ± 0.31 | 69.77 ± 0.21 |
| **Proposed** | **92.97 ± 0.43** | **91.91 ± 0.24** | **87.46 ± 0.13** | **84.75 ± 0.9** | **81.55 ± 0.21** | **79.75 ± 0.92** |

**Table 3:** Mean AUPRC scores (in percentage) for MICU Transfer Prediction on **PHOP** and **MIMIC-IV** Datasets across 3 runs.

| Model | PHOP |  |  | MIMIC-IV |  |  |
| --- | --- | --- | --- | --- | --- | --- |
|  | 1-day | 2-day | 3-day | 1-day | 2-day | 3-day |
| **LR** [22] | 7.52 ± 0.18 | 9.26 ± 0.19 | 6.13 ± 1.47 | 4.27 ± 0.02 | 6.28 ± 0.03 | 4.17 ± 0.04 |
| **FFN** [23] | 9.16 ± 0.31 | 9.91 ± 1.41 | 7.61 ± 0.37 | 6.38 ± 0.11 | 7.05 ± 0.16 | 4.38 ± 0.27 |
| **LSTM** [24] | 9.55 ± 0.38 | 10.26 ± 0.15 | 7.92 ± 0.45 | 9.16 ± 0.17 | 8.45 ± 0.12 | 6.48 ± 0.03 |
| **SGCN** [25] | 12.49 ± 0.13 | 10.91 ± 0.21 | 9.72 ± 0.18 | 11.52 ± 0.37 | 12.64 ± 0.03 | 9.67 ± 0.25 |
| **MT** [26] | 6.82 ± 0.19 | 8.52 ± 0.33 | 5.18 ± 0.02 | 4.13 ± 0.51 | 3.18 ± 0.08 | 3.04 ± 0.10 |
| **HyGCL** [20] | 18.78 ± 1.76 | 17.83 ± 0.11 | 12.71 ± 0.82 | 13.35 ± 0.77 | 14.41 ± 0.45 | 10.05 ± 1.73 |
| **DECent** [27] | 13.16 ± 0.61 | 11.28 ± 0.51 | 9.86 ± 0.39 | 25.63 ± 0.49 | 13.71 ± 0.84 | 9.94 ± 0.69 |
| **HypEHR** [28] | 20.16 ± 0.91 | 13.69 ± 0.58 | 8.55 ± 0.19 | 10.18 ± 0.53 | 8.35 ± 0.06 | 6.38 ± 0.7 |
| **CACHE** [17] | 24.41 ± 0.37 | 15.88 ± 0.72 | 10.61 ± 0.57 | 27.74 ± 0.58 | 10.02 ± 0.15 | 9.02 ± 0.26 |
| **Proposed** | **31.61 ± 0.72** | **19.36 ± 0.15** | **15.58 ± 0.16** | **34.23 ± 0.43** | **17.92 ± 0.64** | **13.13 ± 1.73** |

**Table 4:** AUROC and AUPRC scores (in percentage) for CDI Incidence Prediction on **PHOP** and **MIMIC-IV** Datasets across 3 runs.

| Model | PHOP |  | MIMIC-IV |  |
| --- | --- | --- | --- | --- |
|  | AUROC | AUPRC | AUROC | AUPRC |
| **LR** [22] | 53.25 ± 0.10 | 5.83 ± 0.42 | 66.26 ± 0.81 | 15.83 ± 0.28 |
| **FFN** [23] | 60.30 ± 0.13 | 9.16 ± 0.25 | 71.58 ± 0.55 | 17.47 ± 0.04 |
| **LSTM** [24] | 59.15 ± 0.12 | 9.54 ± 0.13 | 73.26 ± 0.14 | 29.51 ± 0.18 |
| **SGCN** [25] | 71.65 ± 0.04 | 12.59 ± 0.35 | 80.23 ± 0.21 | 48.52 ± 0.16 |
| **MT** [26] | 64.15 ± 0.03 | 9.41 ± 0.02 | 73.94 ± 0.12 | 22.95 ± 0.72 |
| **HyGCL** [20] | 72.38 ± 0.09 | 17.28 ± 0.11 | 85.92 ± 0.24 | 50.79 ± 0.38 |
| **DECent** [27] | 70.83 ± 0.61 | 12.16 ± 0.52 | 77.68 ± 0.10 | 41.61 ± 1.74 |
| **HypEHR** [28] | 67.62 ± 0.45 | 15.17 ± 0.21 | 75.19 ± 0.02 | 38.51 ± 0.06 |
| **CACHE** [17] | 72.18 ± 0.10 | 18.97 ± 0.44 | 79.86 ± 0.39 | 42.75 ± 2.58 |
| **Proposed** | **74.98 ± 0.09** | **21.47 ± 0.14** | **88.32 ± 0.83** | **53.05 ± 0.92** |

### 3.6 Ablation Studies

In this section, we present ablation studies to demonstrate the importance of incorporating domain knowledge and the contributions of each component of our proposed approach. To this end, we conducted both data and model ablation on CDI incidence prediction and 1-day ahead MICU Transfer Prediction tasks on both datasets. We first removed each interaction type, one at a time, then removed different components of our framework and recorded the drop in performance. Figure 2 shows the results for **MIMIC-IV** and the results for **PHOP** are presented in A.2.

**Figure 2.**
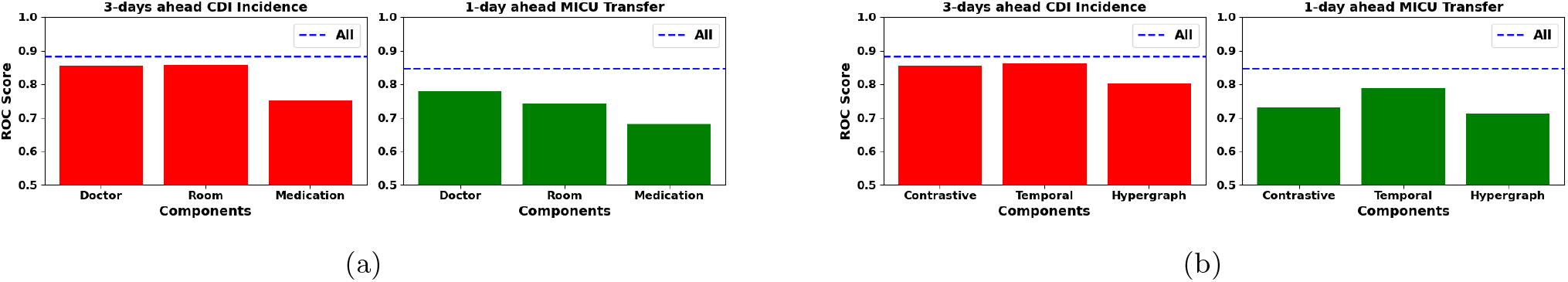
Ablation Studies for **MIMIC-IV** (a) shows the result of removing each type of interaction while (b) shows the result of removing different components of our proposed model.

First, we notice that removal of any interaction leads to a significant drop in performance in all settings indicating that all three interactions are very important for the superior performance of our approach. For **MIMIC-IV**, medication interactions are the dominant type, as their elimination causes the most significant drop in AUROC for both tasks. This is because medication-patient interactions dominate doctor-patient and doctor-room interactions (See Table 1). We also observed that removing doctor interactions and room/unit interactions have similar effects on both CDI Incidence Prediction MICU Transfer Prediction in **MIMIC-IV**. This indicates that these two types of information play roughly equal roles in health risk prediction.

### 3.7 Sensitivity Test

We evaluated the fluctuation in the performance of our framework due to the variation of the hyperparameters and tested performance by varying the latent embedding size by {16, 32, 64] and the learning rate by {0.01, 0.001, 0.0001] . The results for **MIMIC-IV** are in Figure 3 while **PHOP** is in A.3. The results prove the robustness of our framework to the variation of hyperparameters as there is no notable variation in performance by changing either the learning rate or the latent embedding size. Based on the results, a hidden embedding size of 32 and a learning rate of 0.01 were chosen for all our experiments.

**Figure 3.**
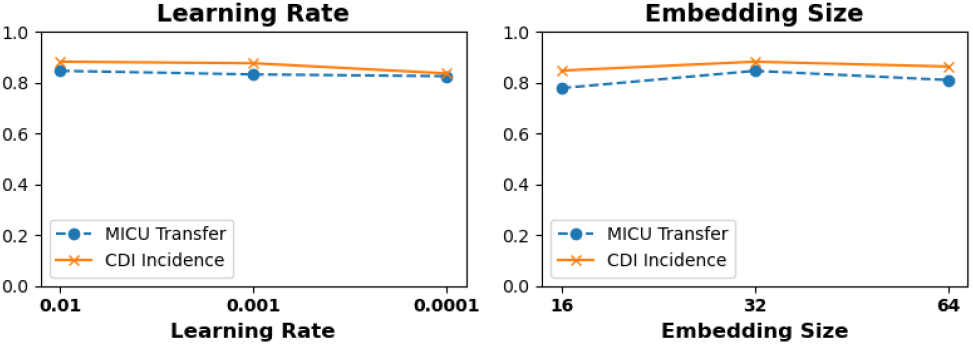
Variation of Learning Rate for CDI Incidence (left) and MICU Transfer (right) for 3-day CDI incidence and 1-day MICU Transfer **MIMIC-IV**

**Figure 4.**
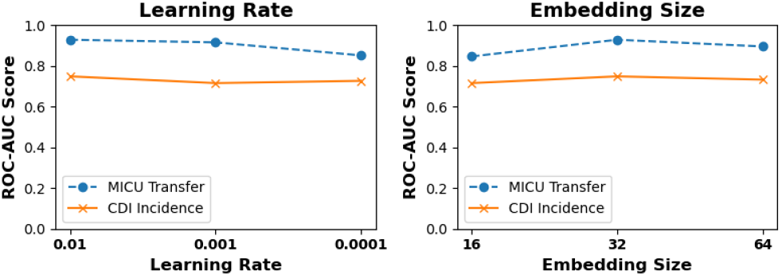
Variation of Learning Rate for CDI Incidence (left) and MICU Transfer (right) for **PHOP**

## 4 RELATED WORKS

### Learning on Heterogenous Graphs and Hypergraphs

Many prior works across other domains use interaction structures for downstream tasks. [34, 35] perform classification on static heterogeneous graphs while [36] considers that the general data contains some elements that have more influence on the whole data than others and [37] uses cooperative learning of spatiotemporal feature embedding for time series prediction. THINK [38] used a hypergraph learning method that captured the hyperbolic properties of time-evolving dynamic hypergraphs. HyperGCN [39] expands hypergraphs to pairwise graphs and samples the relations.

### Health Risk Estimation

Prior works for Healthcare Analytics use patient mobility logs to solve inference problems, such as outbreak detection [40], missing infection [14] and time-series forecasting [41]. Some works use heterogeneous co-evolving networks to learn patient embeddings [15, 27], or utilize the inherent hierarchy present in EHR data [42]. Another direction of active research is to embed medical concepts from EHR data using CNNs [43], or incorporating nonnegativity constraints and structural smoothness [44]. Several works also perform HAI incidence prediction by constructing features [26, 33], or interactions [15, 27]. Overall patient outcome-level prediction is also popular as evidenced by [8, 28, 45, 46].

### Contrastive Learning

Several prior works have explored contrastive learning for graphs and hypergraphs. [47] generates medical code graphs and patient graphs, and leverages contrastive learning to aggregate information. Augmenting hyperedges provides the most numerical gains [48] while [49] generates the same insight for graphs. However, networks provide opportunities to generate augmentations from various levels of structural perspectives [50] while [51] proposes a contrastive loss based on collaborative filtering.

## 5 CONCLUSION

Our proposed contrastive dynamic hypergraph augmentation framework guided by domain knowledge offers a promising solution to the challenges of risk prediction in healthcare. Although recent machine learning approaches have shown promise, they struggle to capture the complexity of heterogeneous interactions leveraged via mobility and fully utilize the domain knowledge, limiting their effectiveness. Our proposed model overcomes the existing limitation by explicitly modeling multifaceted interactions between health entities using a temporally dynamic hypergraph structure and accurately captures the high-order relationships and domain dependencies crucial for precise risk estimation.

## Data Availability

MIMIC-IV Data is publicly available

https://mimic.mit.edu

## A Appendix

### A.1 Further Details about Datasets

#### A.1.1 PHOP

In this data, each patient visit includes a list of diagnoses, a timestamped record of room transfers, physician-performed procedures, and prescription medications. We extracted patient contacts with medications, doctors, and rooms in 2010 between January 1 and March 31. For the period, there were interactions between 6496 unique patients, 575 unique doctors, 686 unique medicines, and 557 rooms.

#### A.1.2 MIMIC-IV

The database contains information on 46,520 patients from 2008 to 2019 and includes demographic information, International Classification of Diseases codes (ICD-9 and ICD-10), hourly vital signs, laboratory tests and microbiological culture results, medication administrations, and survival statistics. All dates in MIMIC-IV are shifted by a factor of years to protect patient information but the sequence of hospital events for every patient is maintained. Compared to MIMIC-III [52], which receives data from heterogeneous sources, MIMIC-IV has more patient data and precise information on procedure events, which are a primary source of clinical information in the ICU, making MIMIC-IV data homogeneous. We extracted patient contacts with medications, doctors, and units between January 1, 2128, and April 4, 2128. Note that the dates in the dataset are shifted from the true values to ensure patient data privacy.

#### A.1.3 Data Split

The split is as follows: (1) **PHOP**: MICU Transfer-1/1/2010-2/7/2010 train, 2/8/2010-2/21/2010 validation, 2/22/2010-3/31/2010 test. CDI Incidence: 1/1/2010-2/20/2010 train, 2/21/2010-3/12/2010 validation, 3/13/2010-3/31/2010 test.(2) **MIMIC-IV**: MICU Transfer-1/1/2128-2/20/2128 train, 2/21/2128-3/11/2128 validation, 3/12/2128-4/4/2128 test. CDI Incidence: 1/1/2128-2/19/2128 train, 2/20/2128-3/1/2128 validation, 3/2/2128-4/4/2128 test.

### A.2 Ablation Study for PHOP

The results of the sensitivity study for **PHOP** are present in Figure 5. Like **MIMIC-IV** medication interactions are the dominant type, as their elimination causes the most significant drop in ROC-AUC for both tasks. Similarly, we also observed that removing doctor interactions and room/unit interactions have similar effects on both CDI Incidence Prediction MICU Transfer Prediction in **PHOP**. When we look at different components being removed, we first note that removal of any component leads to a drop in performance indicating that all components are critical in maintaining the performance of our proposed approach. We performed additional experiments on **PHOP** data for both MICU transfer prediction and CDI prediction where we replaced our semantic knowledge-based augmentations with random augmentations while keeping everything else in our model fixed. We noticed that the performance dropped by up to 4.7% in MICU transfer prediction and by up to 1.5 % in CDI prediction. This result highlights the importance of incorporating semantic knowledge for the prediction of health risks.

**Figure 5.**
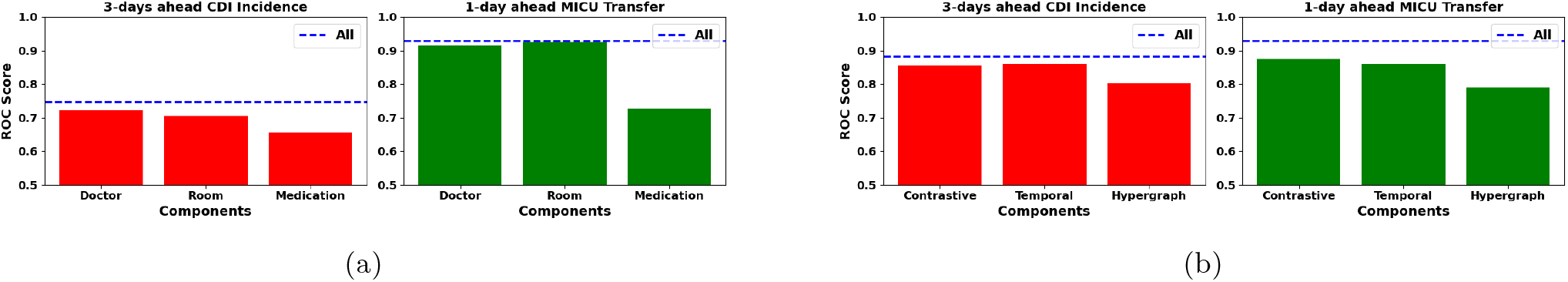
Ablation Studies for **PHOP** (a) shows the result of removing each type of interaction while (b) shows the result of removing different components of our proposed model.

### A.3 Sensitivity Study for PHOP We show the result of varying the parameters for **PHOP** and show them in Figure 4. Notice that the embedding size of 32 gives the best performance for both tasks and the learning rate of 0.01 gives the best performance in this case as well.

## References

[1] C. for Disease Control, P. (CDC et al., “Outbreak of shiga toxin-producing escherichia coli o111 infections associated with a correctional facility dairy-colorado, 2010,” MMWR. Morbidity and mortality weekly report, vol. 61, no. 9, pp. 149–152, 2012.

[2] N. Tomašev, N. Harris, S. Baur, A. Mottram, X. Glorot, J. W. Rae, M. Zielinski, H. Askham, A. Saraiva, V. Magliulo et al., “Use of deep learning to develop continuous-risk models for adverse event prediction from electronic health records,” Nature Protocols, vol. 16, no. 6, pp. 2765–2787, 2021.

[3] P. Saha, R. Sircar, and A. Bose, “Using hospital admission, discharge & transfer (adt) data for predicting readmissions,” Machine Learning with Applications, vol. 5, p. 100055, 2021.

[4] J. Jiang, Y. Wei, Y. Feng, J. Cao, and Y. Gao, “Dynamic hypergraph neural networks.” in IJCAI, 2019, pp. 2635–2641.

[5] J. Luo, M. Ye, C. Xiao, and F. Ma, “Hitanet: Hierarchical time-aware attention networks for risk prediction on electronic health records,” in Proceedings of the 26th ACM SIGKDD International Conference on Knowledge Discovery & Data Mining, 2020, pp. 647– 656.

[6] X. Min, B. Yu, and F. Wang, “Predictive modeling of the hospital readmission risk from patients’ claims data using machine learning: a case study on copd,” Scientific reports, vol. 9, no. 1, p. 2362, 2019.

[7] M. D. Zilberberg, K. Reske, M. Olsen, Y. Yan, and E. R. Dubberke, “Risk factors for recurrent clostridium difficile infection (cdi) hospitalization among hospitalized patients with an initial cdi episode: a retrospective cohort study,” BMC infectious diseases, vol. 14, no. 1, pp. 1–8, 2014.

[8] J. Luo, M. Ye, C. Xiao, and F. Ma, “Hitanet: Hierarchical time-aware attention networks for risk prediction on electronic health records,” in Proceedings of the 26th ACM SIGKDD International Conference on Knowledge Discovery & Data Mining, 2020, pp. 647–656.

[9] Z. Wu, S. Pan, F. Chen, G. Long, C. Zhang, and S. Y. Philip, “A comprehensive survey on graph neural networks,” IEEE transactions on neural networks and learning systems, vol. 32, no. 1, pp. 4–24, 2020.

[10] J. Shang, T. Ma, C. Xiao, and J. Sun, “Pre-training of graph augmented transformers for medication recommendation,” arXiv preprint 1906.00346, 2019.

[11] Y. Shang, Y. Tian, M. Zhou, T. Zhou, K. Lyu, Z. Wang, R. Xin, T. Liang, S. Zhu, and J. Li, “Ehr-oriented knowledge graph system: toward efficient utilization of non-used information buried in routine clinical practice,” IEEE Journal of Biomedical and Health Informatics, vol. 25, no. 7, pp. 2463–2475, 2021.

[12] J. King, V. Patel, E. W. Jamoom, and M. F. Furukawa, “Clinical benefits of electronic health record use: national findings,” Health services research, vol. 49, no. 1pt2, pp. 392–404, 2014.

[13] S. Keyhani, P. L. Hebert, J. S. Ross, A. Federman, C. W. Zhu, and A. L. Siu, “Electronic health record components and the quality of care,” Medical care, pp. 1267–1272, 2008.

[14] P. S. A. B. Jang, H., “Risk-aware temporal cascade reconstruction to detect asymptomatic cases,” Knowledge and Information Systems, vol. 64, p. 3373–3399, 2022.

[15] A. Choudhuri, H. Jang, A. M. Segre, P. M. Polgreen, K. Jha, and B. Adhikari, “Continually-adaptive representation learning framework for time-sensitive healthcare applications,” in Proceedings of the 32nd ACM International Conference on Information and Knowledge Management, ser. CIKM ’23. New York, NY, USA: Association for Computing Machinery, 2023, p. 4538–4544. [Online]. Available: 10.1145/3583780.3615464

[16] Z. Ebnehoseini, M. Tara, M. Meraji, K. Deldar, F. Khoshronezhad, and S. Khoshronezhad, “Usability evaluation of an admission, discharge, and transfer information system: a heuristic evaluation,” Open access Macedonian journal of medical sciences, vol. 6, no. 11, p. 1941, 2018.

[17] R. Xu, Y. Yu, C. Zhang, M. K. Ali, J. C. Ho, and C. Yang, “Counterfactual and factual reasoning over hypergraphs for interpretable clinical predictions on ehr,” in Machine Learning for Health. PMLR, 2022, pp. 259–278.

[18] Y. Zhu, Y. Xu, F. Yu, Q. Liu, S. Wu, and L. Wang, “Graph contrastive learning with adaptive augmentation,” in Proceedings of the Web Conference 2021, ser. WWW ’21. New York, NY, USA: Association for Computing Machinery, 2021, p. 2069–2080. [Online]. Available: 10.1145/3442381.3449802

[19] Y. Qian, C. Zhang, Y. Zhang, Q. Wen, Y. Ye, and C. Zhang, “Co-modality graph contrastive learning for imbalanced node classification,” in Proceedings of the 36th International Conference on Neural Information Processing Systems, ser. NIPS ’22. Red Hook, NY, USA: pnCurran Associates Inc., 2024.

[20] T. Ma, Y. Qian, C. Zhang, and Y. Ye, “Hypergraph contrastive learning for drug trafficking community detection,” in 2023 IEEE International Conference on Data Mining (ICDM), 2023, pp. 1205–1210.

[21] A. Vaswani, N. Shazeer, N. Parmar, J. Uszkoreit, L. Jones, A. N. Gomez, L. Kaiser, and I. Polosukhin, “Attention is all you need,” 2017.

[22] J. Berkson, “Application of the logistic function to bioassay,” Journal of the American statistical association, vol. 39, no. 227, pp. 357–365, 1944.

[23] P. J. Werbos, “Applications of advances in nonlinear sensitivity analysis,” in System Modeling and Optimization: Proceedings of the 10th IFIP Conference New York City, USA, August 31–September 4, 1981. Springer, 2005, pp. 762–770.

[24] S. Hochreiter and J. Schmidhuber, “Long short-term memory,” Neural computation, vol. 9, no. 8, pp. 1735– 1780, 1997.

[25] T. N. Kipf and M. Welling, “Semi-supervised classification with graph convolutional networks,” arXiv preprint 1609.02907, 2016.

[26] J. Oh, M. Makar, C. Fusco, R. McCaffrey, K. Rao, E. E. Ryan, L. Washer, L. R. West, V. B. Young, J. Guttag et al., “A generalizable, data-driven approach to predict daily risk of clostridium difficile infection at two large academic health centers,” infection control & hospital epidemiology, vol. 39, no. 4, pp. 425–433, 2018.

[27] H. Jang, S. Lee, D. H. Hasan, P. M. Polgreen, S. V. Pemmaraju, and B. Adhikari, “Dynamic healthcare embeddings for improving patient care,” in 2022 IEEE/ACM International Conference on Advances in Social Networks Analysis and Mining (ASONAM). IEEE, 2022, pp. 52–59.

[28] R. Xu, M. K. Ali, J. C. Ho, and C. Yang, “Hypergraph transformers for ehr-based clinical predictions,” AMIA Summits on Translational Science Proceedings, vol. 2023, p. 582, 2023.

[29] A. E. Johnson, L. Bulgarelli, L. Shen, A. Gayles, A. Shammout, S. Horng, T. J. Pollard, S. Hao, B. Moody, B. Gow et al., “Mimic-iv, a freely accessible electronic health record dataset,” Scientific data, vol. 10, no. 1, p. 1, 2023.

[30] J. Gao, C. Xiao, Y. Wang, W. Tang, L. M. Glass, and J. Sun, “Stagenet: Stage-aware neural networks for health risk prediction,” in Proceedings of The Web Conference 2020, 2020, pp. 530–540.

[31] CDC. (2023, Mar.) Cdc: https://www.cdc.gov/cdiff/what-is.html. [Online]. Available: CDC:https://www.cdc.gov/cdiff/what-is.html

[32] M. Monsalve, S. Pemmaraju, S. Johnson, and P. M. Polgreen, “Improving risk prediction of clostridium difficile infection using temporal event-pairs,” in 2015 International Conference on Healthcare Informatics, 2015, pp. 140–149.

[33] J. Wiens, J. Guttag, and E. Horvitz, “Patient risk stratification with time-varying parameters: a multitask learning approach,” The Journal of Machine Learning Research, vol. 17, no. 1, pp. 2797–2819, 2016.

[34] X. Sun, H. Yin, B. Liu, H. Chen, J. Cao, Y. Shao, and N. Q. Viet Hung, “Heterogeneous hypergraph embedding for graph classification,” in Proceedings of the 14th ACM International Conference on Web Search and Data Mining, ser. WSDM ’21. New York, NY, USA: Association for Computing Machinery, 2021, p. 725–733. [Online]. Available: 10.1145/3437963.3441835

[35] D. Yang, B. Qu, J. Yang, and P. Cudré-Mauroux, “Lbsn2vec++: Heterogeneous hypergraph embedding for location-based social networks,” IEEE Transactions on Knowledge and Data Engineering, vol. 34, no. 4, pp. 1843–1855, 2022.

[36] X. Kang, X. Li, H. Yao, D. Li, B. Jiang, X. Peng, T. Wu, S. Qi, and L. Dong, “Dynamic hypergraph neural networks based on key hyperedges,” Information Sciences, vol. 616, pp. 37–51, 2022. [Online]. Available: https://www.sciencedirect.com/science/article/pii/S0020025522011264

[37] N. Yin, F. Feng, Z. Luo, X. Zhang, W. Wang, X. Luo, C. Chen, and X.-S. Hua, “Dynamic hypergraph convolutional network,” in 2022 IEEE 38th International Conference on Data Engineering (ICDE), 2022, pp. 1621–1634.

[38] S. Agarwal, R. Sawhney, M. Thakkar, P. Nakov, J. Han, and T. Derr, “Think: Temporal hypergraph hyperbolic network,” in 2022 IEEE International Conference on Data Mining (ICDM), 2022, pp. 849–854.

[39] N. Yadati, M. Nimishakavi, P. Yadav, V. Nitin, A. Louis, and P. Talukdar, “Hypergcn: A new method for training graph convolutional networks on hypergraphs,” in Advances in Neural Information Processing Systems (NeurIPS) 32. Curran Associates, Inc., 2019, pp. 1509–1520.

[40] B. Adhikari, B. Lewis, A. Vullikanti, J. M. Jiménez, and B. A. Prakash, “Fast and near-optimal monitoring for healthcare acquired infection outbreaks,” PLoS CompBio, 2019.

[41] E. Sherman, H. Gurm, U. Balis, S. Owens, and J. Wiens, “Leveraging clinical time-series data for prediction: a cautionary tale,” in AMIA, 2017.

[42] E. Choi, C. Xiao, W. F. Stewart, and J. Sun, “Mime: Multilevel medical embedding of electronic health records for predictive healthcare,” 2018.

[43] Z. Zhu, C. Yin, B. Qian, Y. Cheng, J. Wei, and F. Wang, “Measuring patient similarities via a deep architecture with medical concept embedding,” in IEEE ICDM, 2016.

[44] T. Tran, T. D. Nguyen, D. Phung, and S. Venkatesh, “Learning vector representation of medical objects via EMR-driven nonnegative restricted Boltzmann machines (eNRBM),” J Biomed Inform, 2015.

[45] J. C. Ho, L. R. Staimez, K. V. Narayan, L. Ohno-Machado, R. L. Simpson, and V. S. Hertzberg, “Evaluation of available risk scores to predict multiple cardiovascular complications for patients with type 2 diabetes mellitus using electronic health records,” Computer methods and programs in biomedicine update, vol. 3, p. 100087, 2023.

[46] J. Yi and J. Park, “Hypergraph convolutional recurrent neural network,” in Proceedings of the 26th ACM SIGKDD international conference on knowledge discovery & data mining, 2020, pp. 3366–3376.

[47] D. Cai, C. Sun, M. Song, B. Zhang, S. Hong, and H. Li, Hypergraph Contrastive Learning for Electronic Health Records, pp. 127–135. [Online]. Available: https://epubs.siam.org/doi/abs/10.1137/1.9781611977172.15

[48] T. Wei, Y. You, T. Chen, Y. Shen, J. He, and Z. Wang, “Augmentations in hypergraph contrastive learning: Fabricated and generative,” in Advances in Neural Information Processing Systems, S. Koyejo, S. Mohamed, A. Agarwal, D. Belgrave, K. Cho, and A. Oh, Eds., vol. 35. Curran Associates, Inc., 2022, pp. 1909–1922. [Online]. Available: https://proceedings.neurips.cc/paperfiles/paper/2022/file/0cd1eec0eeaf5ce1bf6d8875a7c1d095-Paper-Conference.pdf

[49] Y. You, T. Chen, Y. Sui, T. Chen, Z. Wang, and Y. Shen, “Graph contrastive learning with augmentations,” in Advances in Neural Information Processing Systems, H. Larochelle, M. Ranzato, R. Hadsell, M. Balcan, H. Lin, Eds., vol. 33. Curran Associates, Inc., 2020, pp. 5812–5823. [Online]. Available: https://proceedings.neurips.cc/paperfiles/paper/2020/file/3fe230348e9a12c13120749e3f9fa4cd-Paper.pdf

[50] D. Lee and K. Shin, “I’m me, we’re us, and i’m us: Tri-directional contrastive learning on hypergraphs,” in Proceedings of the AAAI Conference on Artificial Intelligence, vol. 37, no. 7, 2023, pp. 8456–8464.

[51] L. Xia, C. Huang, Y. Xu, J. Zhao, D. Yin, and J. Huang, “Hypergraph contrastive collaborative filtering,” in Proceedings of the 45th International ACM SIGIR Conference on Research and Development in Information Retrieval, ser. SIGIR ’22. York, NY, USA: Association for Computing Machinery, 2022, p. 70–79. [Online]. Available: 10.1145/3477495.3532058

[52] A. E. Johnson, T. J. Pollard, L. Shen, L.-w. H. Lehman, M. Feng, M. Ghassemi, B. Moody, P. Szolovits, L. Anthony Celi, and R. G. Mark, “Mimic-iii, a freely accessible critical care database,” Scientific data, vol. 3, no. 1, pp. 1–9, 2016.

